# Analytical perturbation reveals hidden instability of biological phenotypes

**DOI:** 10.64898/2026.07.13.26357916

**Authors:** Natalia Piórkowska, Alan Ostromęcki, Grzegorz Franik, Anna Bizoń

## Abstract

**Background:** Unsupervised machine learning has become a cornerstone of computational phenotyping across clinical medicine, genomics, imaging, and multi-omics research. However, phenotype discovery relies on a sequence of analytical decisions – including missing-data handling, preprocessing, dimensionality reduction, clustering methodology, and stochastic initialization – that are rarely evaluated collectively. Although clustering stability has been extensively investigated, the robustness of complete analytical workflows remains largely unexplored.

**Results:** We developed an Analytical Perturbation Framework that systematically quantifies the robustness of phenotype discovery by perturbing complete unsupervised learning workflows rather than individual clustering algorithms. Using a real-world cohort of 1,286 women with polycystic ovary syndrome (PCOS), we generated 116 valid analytical pipelines comprising alternative preprocessing strategies, missing-data handling methods, dimensionality reduction approaches, clustering algorithms, and random initializations. Agreement between independently generated phenotype solutions was consistently low (median Adjusted Rand Index = 0.079), indicating substantial sensitivity of phenotype discovery to routine analytical decisions. Variance decomposition identified preprocessing as the largest contributor to phenotype instability (22.8%), followed by clustering methodology (14.6%), whereas stochastic initialization explained only 3.1% of the observed variability. At the patient level, most individuals exhibited reproducible phenotype assignments (median Patient Robustness Score = 0.719), although a substantial subgroup showed markedly lower assignment stability. Feature perturbation analyses identified follicle-stimulating hormone, anti-thyroglobulin antibodies, anti-thyroid peroxidase antibodies, total testosterone, luteinizing hormone, and androstenedione as the strongest contributors to computational robustness, rather than biological importance. Finally, phenotype solutions demonstrating greater computational robustness also exhibited greater biological coherence during independent validation.

**Availability and implementation:** The analytical framework and reproducible implementation will be made publicly available through a version-controlled repository upon publication.

## 1. Introduction

Computational phenotyping has become one of the fundamental applications of modern bioinformatics and biomedical data science. Advances in high-throughput molecular technologies, electronic health records, medical imaging, and multi-omics integration have enabled increasingly detailed characterization of biological heterogeneity beyond traditional disease classifications. Unsupervised machine learning methods are therefore widely used to identify latent patient subgroups that may represent clinically meaningful phenotypes, molecular disease subtypes, or biologically distinct trajectories relevant to precision medicine [3,7,13–14,21]. Their application now extends across genomics, transcriptomics, metabolomics, single-cell sequencing, spatial biology, and digital health, making computational phenotyping an increasingly important component of translational biomedical research [10,14,21,23].

Despite these advances, computational phenotype discovery remains highly sensitive to methodological decisions embedded within the analytical workflow. Missing-data handling, preprocessing, feature scaling, dimensionality reduction, clustering methodology, and algorithm initialization all modify the geometric representation of the data before phenotype assignment occurs. Because unsupervised learning lacks external ground truth, alternative analytical workflows may generate substantially different phenotype solutions while remaining statistically valid. Consequently, biological conclusions derived from a single computational pipeline may depend as much on analytical design as on the underlying biological structure itself [1–2,5–11].

Several approaches have been proposed to evaluate clustering stability, including bootstrap resampling, consensus clustering, perturbation of input data, and comparisons among clustering algorithms [1,5,9]. These methods provide valuable information regarding algorithmic consistency but typically examine isolated analytical components. In parallel, reproducibility has emerged as a major priority across computational biology, with increasing emphasis on transparent analytical workflows, benchmarking, and reusable bioinformatics pipelines [2,6,10–11]. Nevertheless, current approaches rarely evaluate the cumulative influence of multiple analytical decisions acting simultaneously across complete computational workflows.

This distinction is particularly important because computational phenotyping is inherently a multistage analytical process. Decisions made during preprocessing influence dimensionality reduction, which subsequently affects clustering, patient assignment, downstream interpretation, and biological validation. As a consequence, evaluating only individual clustering algorithms provides an incomplete estimate of computational uncertainty. To our knowledge, no general framework has systematically quantified the robustness of biological phenotypes across complete analytical workflows while simultaneously assessing robustness at the levels of analytical pipelines, individual patients, and individual biomarkers.

In this study, we present an Analytical Perturbation Framework for workflow-level robustness assessment of unsupervised phenotype discovery. Instead of comparing clustering algorithms in isolation, the proposed framework systematically perturbs complete analytical pipelines by varying missing-data handling, preprocessing, dimensionality reduction, clustering methodology, and stochastic initialization. Robustness is subsequently quantified at three complementary levels: agreement between phenotype partitions, reproducibility of phenotype assignment for individual patients, and feature-level contributions to computational robustness. The framework is evaluated using a large real-world cohort of women with polycystic ovary syndrome (PCOS) as a representative case study of a biologically heterogeneous disease. Although PCOS serves as the application domain, the proposed methodology is disease-independent and is intended as a general framework for robustness assessment in computational phenotyping.

## 2. Methods

### 2.1 Study objective

The objective of this study was to evaluate the robustness of biological phenotypes identified using unsupervised machine learning with respect to analytical decisions made throughout the computational workflow. Rather than comparing individual clustering algorithms, we investigated whether phenotype assignment remained stable when complete analytical pipelines were systematically perturbed. To achieve this, we developed an Analytical Perturbation Framework that generated multiple independent unsupervised learning pipelines by varying missing-data handling, preprocessing, dimensionality reduction, clustering methodology, random initialization, and data perturbation strategies. Each pipeline produced an independent phenotype assignment for the same patient cohort.

Phenotype reproducibility was assessed using complementary measures of agreement and stability at the cluster, patient, and feature levels. In particular, we quantified cross-pipeline agreement, patient-level phenotype consistency, and the contribution of individual biomarkers to phenotype stability. The framework was evaluated using a large real-world clinical dataset of women with PCOS. Although PCOS served as the use case, the proposed methodology is disease-agnostic and can be applied to robustness assessment of unsupervised phenotyping in other biomedical datasets.

The analytical framework was evaluated using a retrospective real-world clinical dataset comprising women diagnosed with PCOS at the Department of Endocrinological Gynecology, Medical University of Silesia, Katowice, Poland, between 2018 and 2025. PCOS was diagnosed according to the revised Rotterdam criteria after exclusion of alternative etiologies. Women with incomplete diagnostic evaluation were excluded.

The source database contained 1,331 patients and 224 recorded variables, including demographic characteristics, endocrine hormones, metabolic biomarkers, glucose tolerance measurements, lipid profile, thyroid function tests, hematological parameters, inflammatory markers, and derived clinical indices. Because the database originated from routine clinical practice, laboratory availability differed between patients, resulting in realistic missing-data patterns and heterogeneous feature completeness. The source database additionally included 45 non-PCOS comparison records; after restricting the cohort to confirmed PCOS cases according to the criteria above, the primary analytical cohort used for phenotype discovery comprised 1,286 women.

Patient identifiers were removed before analysis, and all computations were performed on anonymized data. The study was approved by the Bioethical Committee of Wroclaw Medical University (approval No. 254/2021) and conducted in accordance with the Declaration of Helsinki.

The complete computational workflow operated directly on the raw clinical database. Data quality control, variable filtering, preprocessing, feature selection, and all subsequent analytical decisions were performed automatically within a predefined reproducible pipeline.

### 2.3 Analytical framework overview

To evaluate the robustness of unsupervised phenotyping, we developed an Analytical Perturbation Framework that systematically varied the analytical decisions commonly encountered during biomedical data analysis. Instead of relying on a single computational workflow, multiple independent analytical pipelines were generated by combining alternative strategies for missing-data handling, preprocessing, dimensionality reduction, clustering, and data perturbation.

Each pipeline operated on the same patient cohort but represented a distinct sequence of analytical decisions. Consequently, every pipeline produced an independent phenotype assignment. Agreement between these phenotype assignments was subsequently quantified to assess the robustness of the inferred biological structure.

The framework was organized into four consecutive stages: (i) data quality control and preprocessing, (ii) generation of alternative analytical pipelines, (iii) unsupervised phenotype discovery, and (iv) robustness assessment (Figure 1). Robustness was evaluated at three complementary levels. First, cluster-level robustness quantified agreement between complete phenotype partitions generated by different analytical pipelines. Second, patient-level robustness assessed the consistency of phenotype assignment for individual patients across analytical perturbations. Third, feature-level robustness estimated the contribution of individual biomarkers to phenotype stability through systematic feature perturbation analyses.

**Figure 1.**
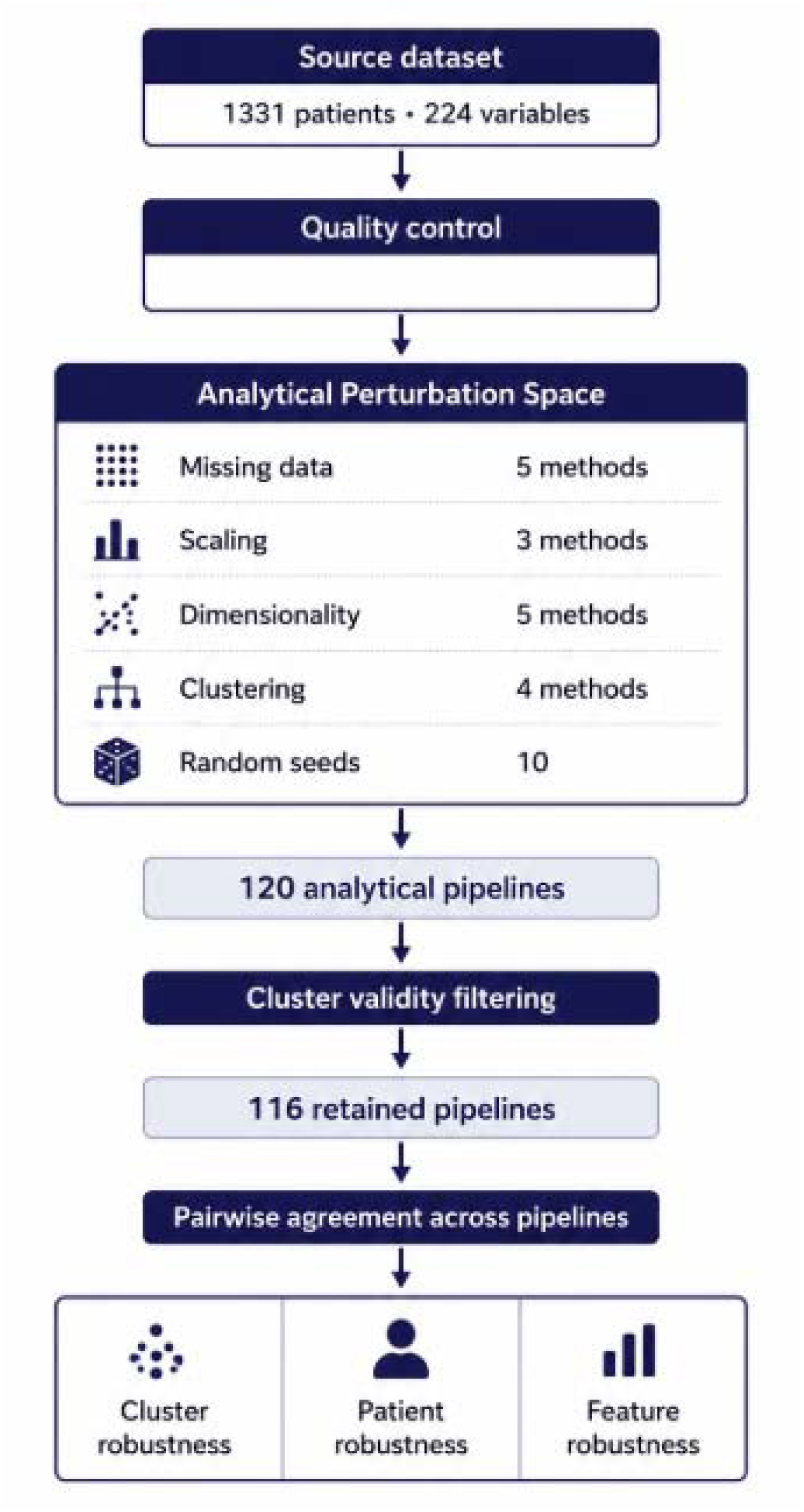
Analytical Perturbation Framework for robustness assessment of unsupervised biological phenotypes.

Unlike conventional benchmarking studies that compare clustering algorithms in isolation, the proposed framework treats the entire analytical workflow as the experimental unit. This design enables evaluation of whether the inferred biological phenotypes represent stable properties of the underlying data or emerge as a consequence of specific analytical choices.

### 2.4 Feature selection and biological spaces

Candidate variables were selected from the original clinical database based on predefined biological relevance and measurement quality. Variables intended for unsupervised phenotyping included routinely acquired endocrine, metabolic, thyroid, hematological, inflammatory, and anthropometric measurements associated with the clinical heterogeneity of PCOS.

Before analysis, variables underwent automated quality control. Features with no recorded observations, excessive missingness, duplicated measurements, technical metadata, or ambiguous biological interpretation were excluded according to prespecified criteria. Highly redundant variables and duplicated laboratory measurements were removed to reduce collinearity while preserving biological interpretability. Derived indices that directly depended on variables used for clustering were excluded from the primary clustering procedure and reserved for downstream biological validation to minimize circularity.

For descriptive purposes, retained variables were grouped into six biological domains: endocrine, metabolic, thyroid, hematological, inflammatory, and anthropometric. These categories were used solely to facilitate biological interpretation of the resulting phenotypes and were not imposed during clustering. All clustering analyses were performed jointly using the complete set of retained features after preprocessing, thereby allowing phenotypic structure to emerge directly from the integrated multidimensional dataset.

The final feature set used for phenotype discovery was determined automatically by the analytical pipeline following completion of quality control and preprocessing, ensuring that feature selection was independent of clustering results and subsequent robustness analyses.

### 2.5 Data quality control

Before phenotype discovery, the source dataset underwent an automated quality-control procedure implemented within the analytical framework. All quality-control operations were predefined and executed without manual intervention to ensure complete reproducibility.

Initially, duplicate patient records and duplicated variables were identified and removed. Variables containing no recorded observations were excluded, whereas variables with excessive missingness were flagged for subsequent evaluation during missing-data handling. Technical metadata, administrative fields, free-text variables, and measurements lacking clear biological interpretation were excluded from further analyses. Continuous variables were inspected for implausible values using predefined physiological ranges and descriptive statistics. Data types were standardized, and laboratory measurements were converted to a consistent numerical format before downstream processing.

To minimize redundancy, highly correlated duplicate measurements originating from repeated laboratory assays or alternative variable encodings were consolidated according to predefined quality-control rules while preserving biologically meaningful information. No manual selection of variables based on clustering performance or downstream analytical results was performed.

Following quality control, an immutable intermediate dataset was generated and used as the exclusive input for all subsequent preprocessing, clustering, and robustness analyses. This procedure ensured that every analytical pipeline originated from an identical quality-controlled dataset, thereby eliminating variability arising from inconsistent data preparation.

### 2.6 Missing data strategies

Missing data handling was treated as an experimental factor rather than a fixed preprocessing step. Instead of selecting a single imputation strategy, the analytical framework systematically evaluated multiple commonly used approaches for handling incomplete clinical data.

Five alternative strategies were considered: (i) complete-case analysis, (ii) median imputation, (iii) k-nearest neighbors (KNN) imputation, (iv) multiple imputation by chained equations using a Bayesian-ridge iterative estimator (MICE), and (v) an iterative ExtraTrees-based imputation procedure (a MissForest-like proxy). Each strategy generated an independent analytical pipeline while all subsequent processing steps remained unchanged.

Variables with no recorded observations were removed during quality control prior to imputation. Missing-value handling was subsequently performed independently within each analytical pipeline to prevent information leakage between alternative analytical workflows. No imputed dataset was reused across pipelines.

Treating missing-data handling as a perturbation factor enabled direct evaluation of its influence on phenotype discovery. Agreement between phenotype assignments generated using different imputation strategies was quantified together with all remaining analytical perturbations as part of the overall robustness assessment.

### 2.7 Preprocessing strategies

Data preprocessing was incorporated into the analytical perturbation framework as an independent source of methodological variability. Rather than applying a single preprocessing protocol, alternative preprocessing strategies were evaluated across separate analytical pipelines to assess their influence on phenotype discovery.

Continuous variables were standardized using one of three commonly applied scaling approaches: z-score standardization, robust scaling, or quantile transformation. Each preprocessing strategy was applied independently within the corresponding analytical pipeline after completion of missing-data handling and before dimensionality reduction.

To avoid information leakage, preprocessing parameters were estimated separately for each analytical pipeline and were never shared across alternative workflows. Consequently, every pipeline represented an independent sequence of preprocessing decisions prior to phenotype discovery.

Preprocessing methods were selected because they represent widely used normalization strategies in biomedical machine learning and differ substantially in their sensitivity to skewed distributions, outliers, and heterogeneous measurement scales. Their inclusion enabled direct evaluation of whether biologically inferred phenotypes remained stable despite reasonable variation in data normalization procedures.

### 2.8 Dimensionality reduction

Dimensionality reduction was evaluated as an additional source of analytical variability within the proposed framework. Instead of adopting a single transformation method, multiple approaches were incorporated into independent analytical pipelines to assess their influence on phenotype discovery.

Five alternative strategies were considered: no dimensionality reduction, principal component analysis (PCA) retained at 70%, 80%, or 90% cumulative explained variance, and independent component analysis (ICA). Each dimensionality reduction method was fitted independently within its corresponding analytical pipeline following preprocessing and before clustering. Transformation parameters were estimated exclusively from the data available within the respective pipeline to ensure complete analytical independence.

Including multiple dimensionality reduction strategies enabled direct assessment of whether the inferred biological phenotypes represented stable properties of the underlying clinical data or were influenced by the choice of latent feature representation.

### 2.9 Clustering algorithms

Unsupervised phenotype discovery was performed using multiple clustering algorithms representing complementary assumptions regarding cluster geometry and data structure. Rather than identifying a single optimal method, clustering algorithms were incorporated as analytical perturbation factors within the proposed framework.

Four clustering approaches were evaluated: k-means, diagonal-covariance Gaussian mixture models (GMM), Ward-linkage agglomerative hierarchical clustering, and a repeated k-means ensemble consensus procedure. Together, these algorithms encompass centroid-based, probabilistic, hierarchical, and ensemble clustering paradigms commonly applied in biomedical data analysis.

Each clustering algorithm was applied independently to the output of the corresponding preprocessing and dimensionality reduction pipeline. Hyperparameters were predefined before analysis and were not optimized according to clustering performance or biological outcomes. The number of clusters was fixed at two for all analytical pipelines, consistent with the primary two-phenotype structure reported in prior PCOS clustering literature. Random initialization was repeated across multiple independent seeds for stochastic algorithms to capture variability arising from algorithm initialization.

To avoid biologically implausible solutions, clustering results were subjected to predefined quality-control criteria. Solutions containing extremely small clusters or failing to satisfy minimum cluster-size requirements were excluded from downstream robustness analyses. This filtering procedure prevented unstable partitions generated by algorithmic artifacts from influencing the assessment of phenotype reproducibility.

The objective of the clustering stage was not to identify the best-performing algorithm but to evaluate whether biologically meaningful phenotypes remained reproducible across fundamentally different clustering paradigms.

### 2.10 Construction of analytical perturbation pipelines

The proposed framework was based on the generation of multiple independent analytical pipelines, each representing a distinct combination of methodological decisions commonly encountered during unsupervised biomedical data analysis. Rather than modifying a single analytical component, complete computational workflows were systematically perturbed to evaluate the robustness of phenotype discovery under realistic analytical variability.

Each analytical pipeline consisted of five consecutive stages: missing-data handling, data preprocessing, dimensionality reduction, clustering, and robustness evaluation. Alternative methods available at each stage were combined according to a predefined experimental design established before analysis. Consequently, every pipeline represented an independent realization of the complete phenotype discovery process applied to the same patient cohort.

To ensure reproducibility, all analytical decisions, random seeds, algorithm parameters, and quality-control criteria were specified before execution and applied consistently across all pipelines. No analytical pathway was modified on the basis of intermediate clustering results or biological interpretation. Pipelines producing degenerate clustering solutions, including partitions containing clusters below the predefined minimum size threshold, were excluded from downstream analyses according to prespecified quality-control rules.

Each valid pipeline generated an independent phenotype assignment. Agreement between these phenotype assignments formed the basis of the subsequent robustness analyses, enabling quantification of phenotype stability at the cluster, patient, and feature levels.

A schematic overview of the analytical perturbation framework is presented in Figure 1, whereas the complete list of evaluated pipeline configurations and algorithmic parameters is provided in the Supplementary Methods.

### 2.11 Cluster validity filtering

Because robustness estimates may be distorted by algorithmically degenerate clustering solutions, all phenotype partitions underwent automated validity assessment before inclusion in downstream analyses. Cluster validity filtering was applied uniformly across all analytical pipelines using predefined criteria established before execution.

Partitions containing clusters below the predefined minimum cluster-size threshold or exhibiting insufficient representation of the study population were excluded from further analyses. This procedure prevented unstable solutions resulting from algorithmic artifacts, random initialization, or over-fragmentation of the data from influencing robustness estimates.

Only phenotype partitions satisfying all validity criteria were retained for subsequent evaluation of cluster-, patient-, and feature-level robustness. Consequently, robustness metrics were calculated exclusively from biologically interpretable and computationally stable clustering solutions.

Cluster validity filtering was performed independently of biological interpretation and without reference to downstream robustness metrics, ensuring that exclusion decisions were based solely on predefined quality-control criteria.

### 2.12 Phenotype robustness metrics

Phenotype robustness was quantified by measuring the agreement between phenotype assignments generated by independent analytical pipelines. Rather than relying on a single similarity measure, robustness was evaluated using a complementary set of partition comparison metrics capturing different aspects of clustering agreement.

Cluster-level agreement between analytical pipelines was assessed using the Adjusted Rand Index (ARI), Normalized Mutual Information (NMI), Variation of Information (VI), and pairwise co-assignment measures. Together, these metrics quantify partition similarity while accounting for differences in cluster size, label permutation, and information content.

To evaluate robustness across the entire analytical framework, agreement was calculated for all valid pairwise combinations of analytical pipelines. This approach enabled estimation of phenotype stability independently of any individual clustering algorithm or preprocessing strategy.

Robustness assessment was performed at three complementary levels. Cluster-level robustness quantified the reproducibility of complete phenotype partitions. Patient-level robustness evaluated the consistency of phenotype assignment across analytical perturbations for individual patients. Feature-level robustness estimated the contribution of individual variables to phenotype stability through systematic feature perturbation analyses.

Because robustness rather than clustering performance represented the primary outcome of the study, all subsequent analyses focused on agreement between independently generated phenotype assignments instead of conventional internal clustering quality measures.

### 2.13 Patient-level robustness

In addition to evaluating agreement between complete phenotype partitions, robustness was assessed at the individual patient level. This analysis was designed to determine whether each patient was assigned consistently to the same biological phenotype across independent analytical pipelines.

For every patient, a label-invariant Patient Robustness Score (PRS) was calculated from pairwise co-assignment: for each pair of analytical pipelines, two patients were scored as concordant if they were placed in the same phenotype together in both pipelines, regardless of which numerical label that phenotype received. The PRS for a given patient was defined as the proportion of concordant co-assignments with their consensus neighborhood across all valid pipeline pairs. Because concordance is evaluated on shared cluster membership rather than on label identity, the PRS is unaffected by label switching between independently fitted pipelines. Higher PRS values indicate stable phenotype assignment regardless of analytical decisions, whereas lower values identify patients whose phenotype membership changes substantially under analytical perturbation.

To further characterize assignment uncertainty, complementary measures of assignment entropy and phenotype persistence were calculated. Assignment entropy quantified the variability of phenotype membership across analytical pipelines, while phenotype persistence represented the proportion of valid pipelines assigning a patient to the dominant phenotype. Together, these measures distinguish patients exhibiting stable biological phenotypes from individuals located near phenotype boundaries.

Patient-level robustness was evaluated independently of clinical outcomes and was used exclusively to characterize the stability of phenotype assignment. Summary distributions of patient robustness metrics were subsequently compared across analytical pipelines to identify regions of stable and unstable phenotypic organization within the study cohort.

### 2.14 Feature robustness analysis

To identify variables contributing to phenotype stability, feature robustness was evaluated using a systematic perturbation analysis. Individual variables were temporarily removed from the analytical workflow while all remaining processing steps were preserved. The resulting phenotype assignments were subsequently compared with the corresponding reference pipeline to quantify the influence of each variable on phenotype reproducibility.

Feature perturbation was performed independently across valid analytical pipelines to reduce dependence on individual preprocessing strategies or clustering algorithms. For each variable, robustness estimates were aggregated across repeated perturbation analyses, providing a global measure of its contribution to phenotype stability rather than to any specific clustering solution.

Variables whose removal consistently reduced agreement between phenotype assignments were interpreted as stabilizing components of the phenotypic structure. Conversely, variables producing minimal changes in phenotype assignment were considered to have limited influence on phenotype robustness within the evaluated analytical framework.

Feature robustness analysis was performed exclusively to characterize the stability of unsupervised phenotyping and should not be interpreted as a measure of biological importance or disease causality. Instead, the analysis quantifies the extent to which individual variables contribute to the reproducibility of phenotype discovery under analytical perturbation.

### 2.15 Biological validation

Biological validation was performed to determine whether analytically robust phenotype assignments were also biologically meaningful. Rather than validating individual clustering algorithms, validation was conducted at the level of phenotype robustness generated by the analytical perturbation framework.

To avoid circularity, biological validation was based exclusively on variables that were not directly involved in the corresponding clustering analysis whenever possible. Clinical characteristics, laboratory measurements, and derived indices reserved for downstream evaluation were compared across robust phenotype assignments to assess their biological consistency.

Patients were additionally stratified according to their PRS. Clinical and laboratory characteristics of patients exhibiting highly stable phenotype assignments were compared with those showing low assignment stability. This analysis enabled evaluation of whether analytically stable phenotypes corresponded to more coherent biological profiles.

Variables derived directly from clustering features (e.g., composite clinical indices) were considered exploratory and interpreted separately from the primary validation analyses. Consequently, the principal biological validation relied on independent measurements whenever available, thereby minimizing circularity between phenotype discovery and phenotype evaluation.

The objective of biological validation was not to maximize statistical separation between phenotype groups but to determine whether robustness under analytical perturbation was associated with increased biological consistency of the resulting phenotypes.

### 2.16 Statistical analysis

Descriptive statistics were used to summarize the characteristics of the study population and the distributions of robustness metrics. Continuous variables are reported as mean ± standard deviation (SD) or median with interquartile range (IQR), depending on data distribution, whereas categorical variables are presented as counts and percentages.

Comparisons between phenotype groups or robustness strata were performed using non-parametric statistical tests appropriate for the distribution of the biological validation variables. Group-level differences across the reference phenotype partition were assessed using the Kruskal–Wallis H test, pairwise comparisons between low– and high-robustness patient strata were assessed using the Mann–Whitney U test, and associations between continuous biomarkers and patient co-assignment robustness were assessed using Spearman rank correlation. When multiple comparisons were performed, all resulting p-values were adjusted using the Benjamini– Hochberg false discovery rate (FDR) procedure.

Robustness metrics were summarized using descriptive statistics and empirical distributions across all valid analytical pipelines. Confidence intervals for selected robustness estimates were obtained by bootstrap resampling where applicable. No hypothesis testing was performed to compare alternative analytical pipelines, as the primary objective was to characterize robustness rather than identify a superior analytical strategy.

All statistical tests were two-sided, and a *P* value < 0.05 was considered statistically significant unless otherwise specified. Statistical analyses were predefined before execution of the analytical framework.

### 2.17 Software and reproducibility

All analyses were performed using Python 3 within a fully reproducible computational framework developed in Google Colaboratory. The analytical workflow was implemented as a series of independent notebook-based modules covering data quality control, preprocessing, dimensionality reduction, clustering, robustness assessment, biological validation, and automated figure generation.

To ensure complete reproducibility, all analytical decisions, algorithm parameters, random seeds, and quality-control criteria were predefined before execution. Each analytical pipeline was executed independently from an identical quality-controlled input dataset, and intermediate results were stored after every processing stage to provide full traceability of the computational workflow.

The framework automatically generated reproducibility reports, including software versions, package dependencies, execution parameters, random seeds, and metadata required to reproduce the complete analysis. Data processing was deterministic whenever possible, whereas stochastic procedures were controlled using predefined random initialization.

The complete analytical framework, together with documentation, example input data, and instructions for reproducing the analyses, will be made publicly available through a version-controlled repository upon publication. The exact software versions used for the analyses are provided in the accompanying repository and Supplementary Materials.

## 3. Results

### 3.1 Cohort characteristics and analytical pipeline generation

The analytical perturbation framework was applied to a real-world clinical cohort comprising 1,286 women with confirmed PCOS, derived from an original database containing 1,331 patient records collected during routine clinical care. Following automated quality control, duplicate removal, variable filtering, and predefined preprocessing procedures, all analytical workflows were initialized from an identical quality-controlled dataset, ensuring that subsequent differences between phenotype assignments originated exclusively from analytical perturbations rather than inconsistencies in data preparation.

The framework systematically combined alternative analytical decisions at each stage of the unsupervised learning workflow, including missing-data handling, data preprocessing, dimensionality reduction, clustering methodology, and random initialization. This combinatorial design generated a large collection of independent analytical pipelines, each representing a distinct but methodologically valid implementation of the complete phenotype discovery process (Figure 1).

Application of predefined cluster validity criteria excluded analytical pipelines producing degenerate clustering solutions, such as partitions containing excessively small clusters or failing to satisfy minimum cluster-size requirements. After automated quality-control filtering, 116 independent analytical pipelines were retained for downstream robustness analyses. Each retained pipeline generated an independent phenotype assignment for the same patient cohort, providing the basis for subsequent comparisons of phenotype reproducibility across alternative analytical workflows.

The resulting collection of validated analytical pipelines constituted the experimental foundation for all subsequent robustness analyses presented in this study. Rather than evaluating a single clustering solution, the proposed framework enabled systematic assessment of phenotype reproducibility across a broad spectrum of realistic analytical decisions (Table 1).

**Table 1.** Characteristics of the study cohort and analytical perturbation framework.

| Component | Value | Details |
| --- | --- | --- |
| Source clinical database | 1331 | 224 recorded variables before primary population filtering |
| Confirmed PCOS cohort | 1286 | Primary cohort retained for phenotype discovery |
| Phenotyping variables | 26 | Core endocrine, metabolic, thyroid, hematological and inflammatory features (28 candidate variables screened; 2 excluded for excessive missingness) \| |
| Missing-data strategies | 5 | Complete-case, median, KNN, MICE, MissForest-like iterative imputation |
| Scaling strategies | 3 | Z-score, robust scaling, quantile transformation |
| Dimensionality-reduction strategies | 5 | None, PCA 70%, PCA 80%, PCA 90%, ICA |
| Clustering algorithms | 4 | K-means, diagonal-covariance GMM, Ward hierarchical clustering, consensus clustering |
| Random seeds | 5 | Seeds 0–4 in the frozen MANUSCRIPT_FINAL run |
| Phenotype number | 2 | Fixed for all analytical pipelines |
| Analytical pipelines generated | 120 | Balanced representative perturbation design |
| Pipelines excluded by cluster-validity QC | 4 | Degenerate solutions failing predefined minimum cluster-size criteria |
| Valid pipelines retained | 116 | Included in downstream robustness analyses |

### 3.2 Global phenotype robustness is low across analytical perturbations

To evaluate the overall reproducibility of unsupervised phenotype discovery, agreement between phenotype assignments generated by the 116 valid analytical pipelines was quantified using complementary partition similarity metrics (Figure 2). Despite all pipelines operating on the identical quality-controlled patient cohort, substantial variability in phenotype assignment was observed across alternative analytical workflows.

**Figure 2.**
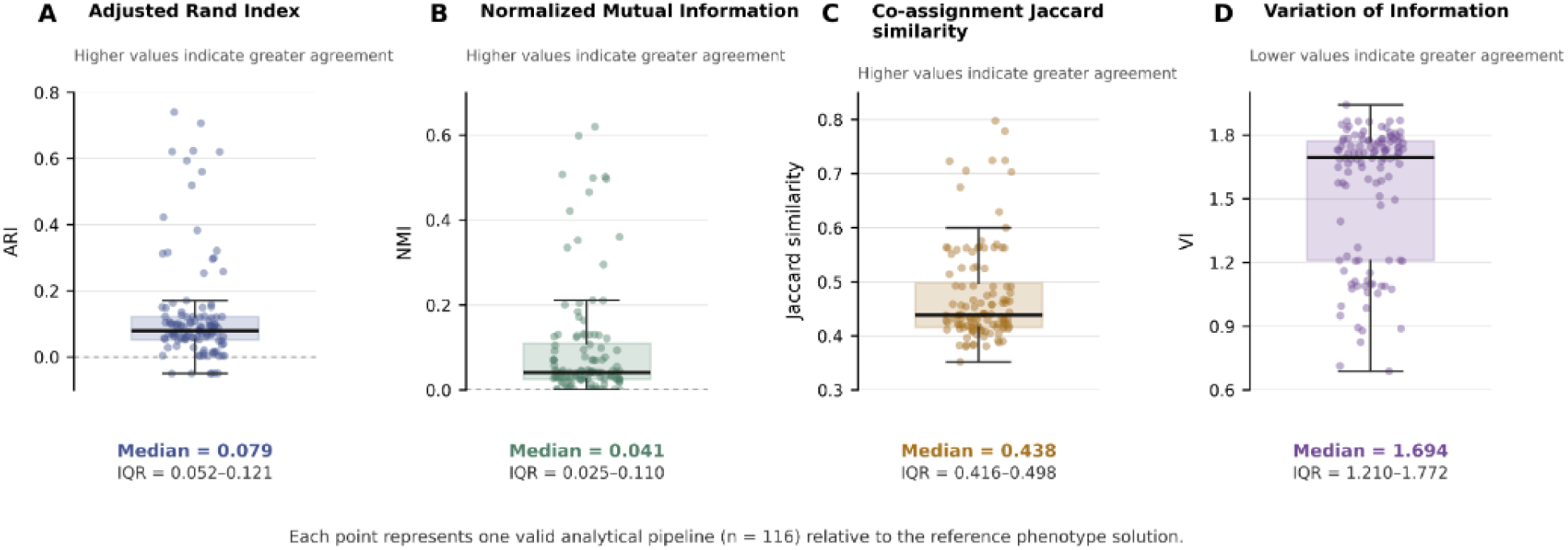
Global robustness of phenotype assignments across analytical perturbations. *Distribution of global agreement metrics calculated across the 116 valid analytical pipelines relative to the reference phenotype solution. Shown are the distributions of the Adjusted Rand Index (ARI), Normalized Mutual Information (NMI), co-assignment Jaccard similarity, and Variation of Information (VI). Despite identical input data, agreement between independently generated phenotype assignments remained consistently low, demonstrating substantial sensitivity of unsupervised phenotype discovery to realistic analytical perturbations. Horizontal lines represent median values and interquartile ranges*.

Agreement with the reference phenotype solution was consistently low across all evaluated robustness metrics. The median ARI was 0.079 (IQR 0.052–0.121), indicating that relatively small modifications of the analytical workflow frequently altered phenotype membership. Similarly, the median NMI reached only 0.041, demonstrating limited preservation of the overall clustering structure across analytical perturbations. Co-assignment similarity remained moderate, with a median Jaccard index of 0.438, whereas the median Variation of Information (VI) reached 1.69, indicating substantial information divergence between independently generated phenotype partitions.

Importantly, the observed variability was not restricted to a single analytical component but emerged despite all pipelines being derived from the same quality-controlled dataset and differing only in commonly accepted analytical decisions, including missing-data handling, preprocessing, dimensionality reduction, clustering methodology, and random initialization. The consistency of this observation across multiple complementary agreement measures indicates that phenotype instability represents a global property of the analytical process rather than an artifact of a specific robustness metric.

Collectively, these findings demonstrate that unsupervised biological phenotypes identified in the present cohort are highly sensitive to realistic analytical perturbations. Rather than representing invariant biological structures, phenotype assignments varied substantially across equally justifiable analytical workflows, highlighting the necessity of explicitly evaluating robustness before interpreting computationally derived biological phenotypes.

### 3.3 Pairwise comparison reveals widespread disagreement between analytical workflows

To assess phenotype reproducibility independently of any single reference solution, all valid analytical pipelines were compared pairwise using the ARI. The 116 retained pipelines yielded 6,670 unique pairwise comparisons, providing a comprehensive assessment of agreement across the complete analytical perturbation space (Figure 3).

**Figure 3.**
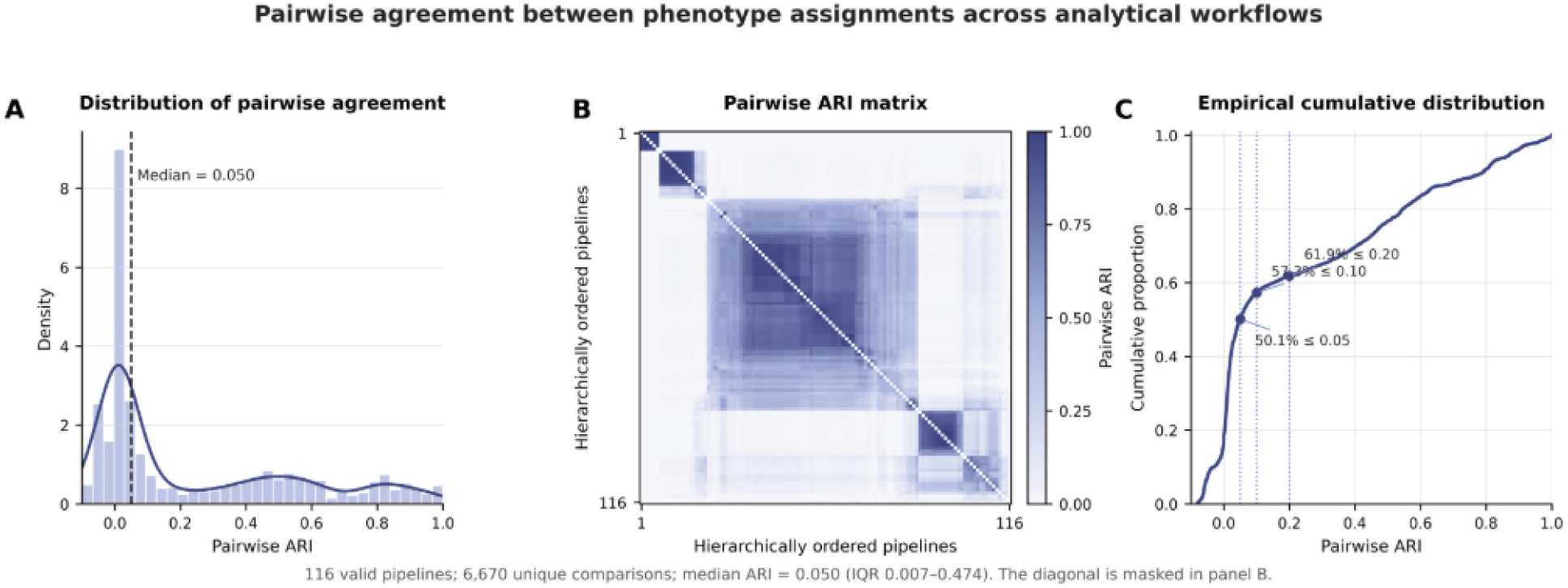
Pairwise agreement between phenotype assignments generated by alternative analytical workflows. *Pairwise Adjusted Rand Index agreement among the 116 valid analytical pipelines. **(A)** Heatmap of th complete pairwise ARI matrix after average-linkage hierarchical ordering for visualization. The diagonal was omitted because it represents trivial self-comparisons. Darker values indicate greater similarity between phenotype partitions. **(B)** Empirical distribution of the 6,670 unique pairwise comparisons. The dashed line indicates the median pairwise ARI of 0.050. The predominance of values close to zero, together with isolated regions of higher agreement, demonstrates widespread disagreement between alternative analytical workflows*.

Pairwise agreement between independently generated phenotype partitions was generally very low. The median pairwise ARI was only 0.049, indicating that most analytical workflows produced substantially different patient assignments despite being applied to the same quality-controlled cohort. The predominance of low ARI values across the comparison matrix demonstrates that phenotype instability was not limited to pipelines that differed strongly from the predefined reference solution. Instead, disagreement was widespread across the full set of alternative workflows.

The pairwise ARI heatmap revealed only a small number of localized regions with high agreement. These regions represented specific combinations of analytical decisions that generated similar phenotype partitions, whereas most pipeline pairs showed limited correspondence. No broad block of consistently high agreement was observed across the perturbation space, suggesting that reproducibility was restricted to isolated workflow configurations rather than representing a general property of the inferred phenotype structure.

These results provide a reference-independent confirmation of global phenotype instability. The very low median pairwise ARI demonstrates that different, methodologically plausible analytical workflows frequently identified distinct partitions of the same patient population. Consequently, the apparent phenotype structure obtained from any individual pipeline cannot be assumed to represent a unique or analytically invariant organization of the cohort.

### 3.4 Preprocessing decisions dominate phenotype instability

To identify which analytical decisions contributed most to phenotype instability, we decomposed the observed variability in phenotype agreement into the individual components of the analytical perturbation framework (Figure 4). This analysis quantified the relative contribution of each methodological choice to the overall variability of phenotype assignments across the complete analytical perturbation space.

**Figure 4.**
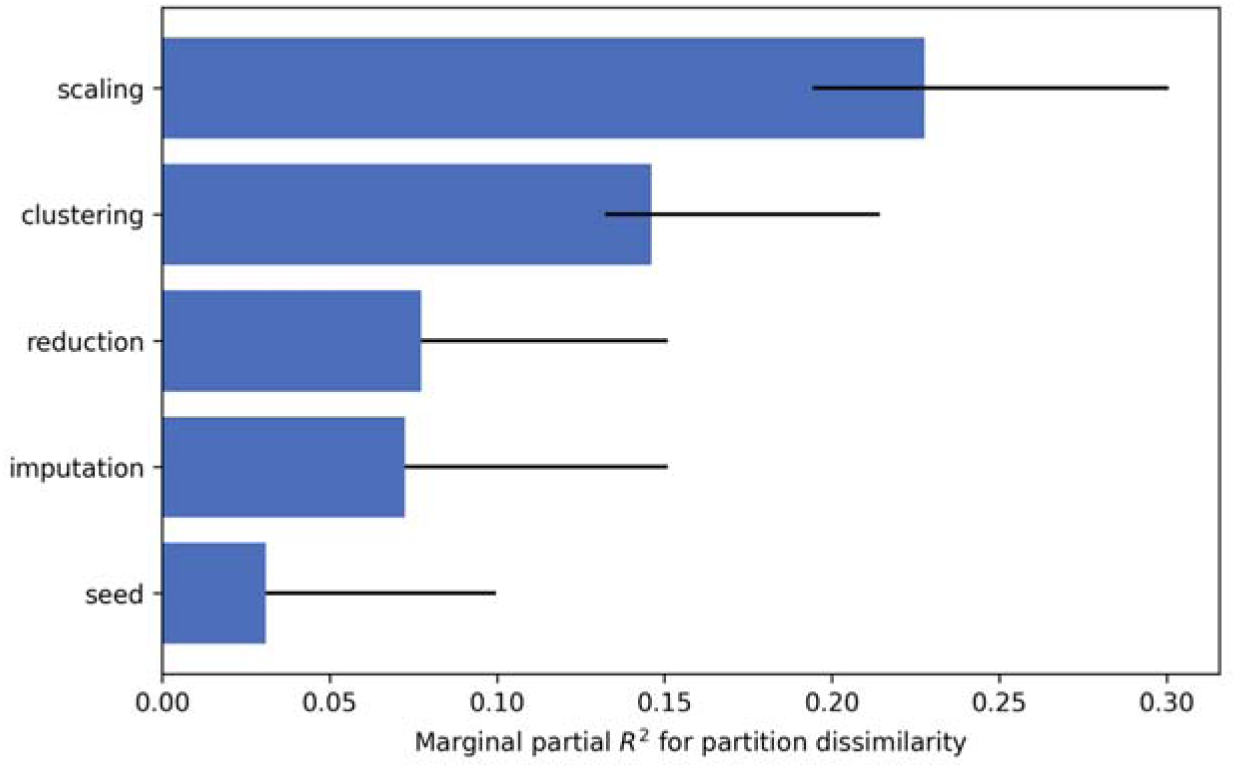
Relative contribution of analytical decisions to phenotype instability. *Relative contribution of individual analytical components to the explainable variability of phenotyp assignments estimated using distance-based variance decomposition. Horizontal bars represent marginal partial R², and black lines indicate 95% bootstrap confidence intervals. Scaling strategy accounted for the largest proportion of phenotype variability (22.8%), followed by the clustering algorithm (14.6%), dimensionality reduction (7.7%), missing-data handling (7.3%), and random initialization (3.1%). These findings indicate that phenotype instability is driven predominantly by deterministic analytical choices rather than stochastic optimization*.

Variance decomposition demonstrated that data preprocessing was the dominant source of phenotype variability, accounting for 22.8% of the explainable variance in phenotype agreement. The choice of clustering algorithm represented the second largest contributor (14.6%), indicating that both data transformation and cluster identification substantially influenced the resulting phenotype structure.

In contrast, the effects of dimensionality reduction and missing-data handling were considerably smaller, explaining 7.7% and 7.3% of the variance, respectively. Although these analytical steps measurably affected phenotype reproducibility, their influence was markedly lower than that of preprocessing and clustering. Notably, random initialization accounted for only 3.1% of the observed variability, demonstrating that stochastic optimization contributed minimally to phenotype instability.

The relative importance of these analytical components indicates that phenotype reproducibility is determined primarily by deterministic methodological choices rather than by random variation during model fitting. In particular, the dominant contribution of preprocessing suggests that transformations applied before clustering exert a greater influence on the final phenotype structure than the stochastic behavior of the clustering algorithms themselves.

Collectively, these findings demonstrate that analytical instability originates predominantly from methodological decisions embedded within the computational workflow. Consequently, improving phenotype reproducibility is unlikely to be achieved through repeated random initialization alone but instead requires explicit evaluation of preprocessing strategies and other key analytical choices during unsupervised phenotype discovery.

### 3.5 Patient-level robustness identifies stable and unstable individuals

Although global agreement between analytical workflows was generally low, robustness varied substantially across individual patients. To quantify phenotype stability at the subject level, we calculated the PRS, defined as the proportion of analytical pipelines assigning a given patient to the same phenotype independently of phenotype label permutation (Figure 5).

**Figure 5.**
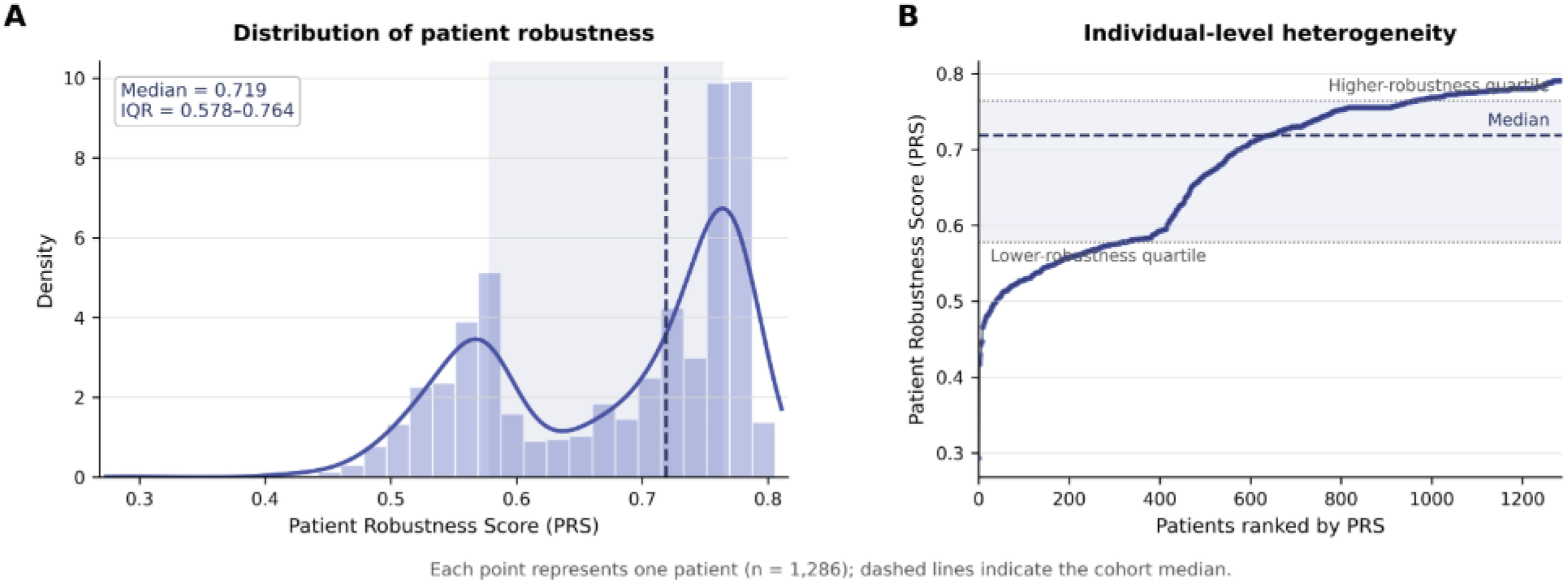
Distribution of patient-level phenotype robustness. *(A) Distribution of the label-invariant Patient Robustness Score (PRS) across 1,286 patients. The dashed vertical line indicates the cohort median, and the shaded region represents the interquartile range. The median PRS was 0.719 (IQR 0.578–0.764). (B) Individual PRS values ordered from lowest to highest, illustrating substantial heterogeneity in assignment stability across the cohort. Horizontal lines indicate the median and quartile boundaries. Although many patients exhibited highly reproducible phenotype assignments, a distinct subset showed lower robustness and greater sensitivity to analytical perturbations*.

The overall distribution of PRS values demonstrated that many individuals were assigned consistently across alternative analytical workflows. The median PRS was 0.719 (IQR 0.578– 0.764), indicating that most patients exhibited relatively stable phenotype membership despite the pronounced instability observed at the global clustering level.

Nevertheless, the distribution revealed considerable heterogeneity in patient-level robustness. While a substantial proportion of individuals showed consistently high PRS values, a sizable intermediate group displayed only moderate assignment stability. These patients frequently changed phenotype membership across equally plausible analytical workflows, suggesting that they occupy transitional regions of the multidimensional biological space rather than belonging unequivocally to a single phenotype.

Importantly, only a small fraction of patients exhibited very low robustness scores, whereas highly robust assignments predominated at the upper end of the distribution. These findings indicate that analytical instability is not uniformly distributed across the cohort but is concentrated in a subset of biologically ambiguous individuals.

Overall, patient-level robustness provides complementary information beyond conventional clustering metrics by identifying which phenotype assignments remain reproducible across analytical perturbations and which should be interpreted with greater caution. Rather than treating all phenotype assignments as equally reliable, the Patient Robustness Score enables confidence to be quantified at the level of individual patients.

### 3.6 Feature perturbation identifies biomarkers stabilizing phenotype discovery

To determine which variables contributed most to the reproducibility of phenotype discovery, we performed systematic feature perturbation analyses in which individual biomarkers were removed from the analytical workflow and the resulting phenotype assignments were compared with those obtained from the corresponding unperturbed pipelines (Figure 6). This approach quantified the contribution of each feature to computational robustness rather than its biological or causal importance.

**Figure 6.**
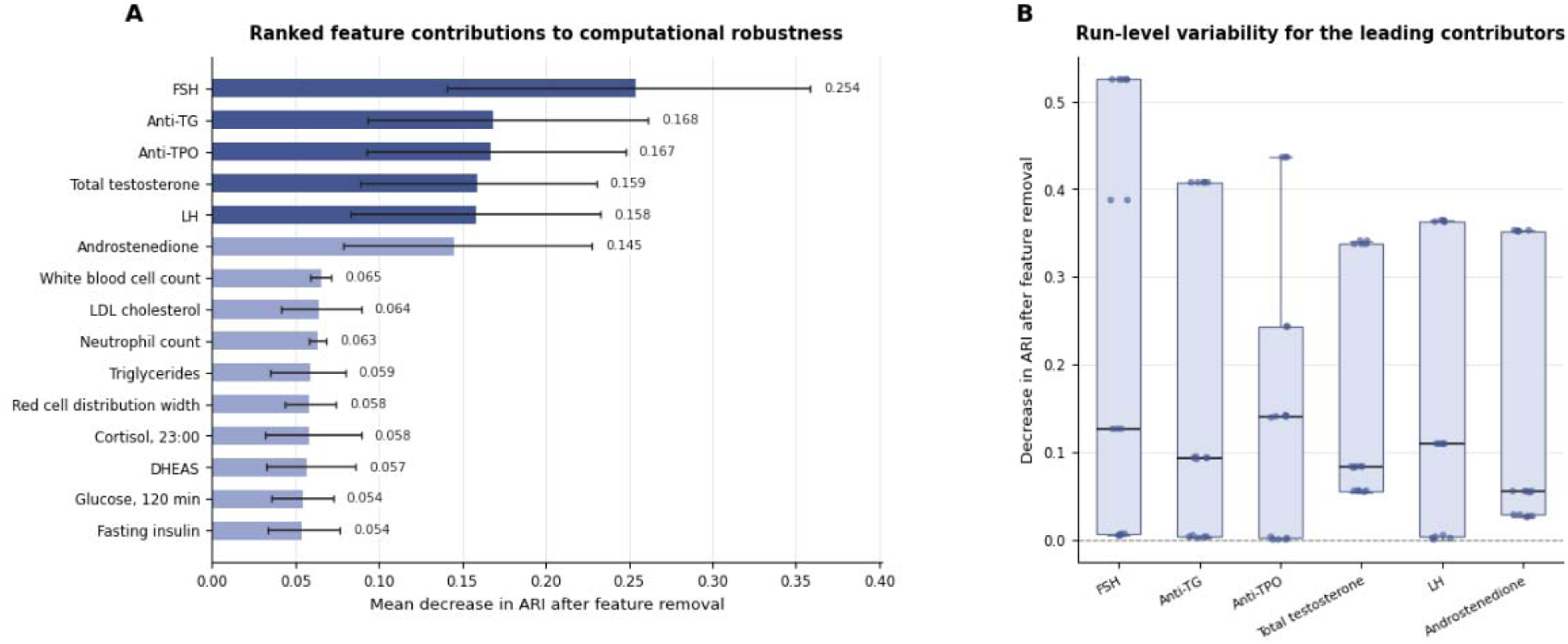
Feature-level contributions to the robustness of phenotype discovery. *(A) Ranked feature-level contributions estimated using systematic leave-one-feature-out perturbation. Bars represent the mean decrease in Adjusted Rand Index after removal of each feature, and horizontal intervals indicate 95% bootstrap confidence intervals. Larger values indicate a greater contribution to the computational robustness of phenotype discovery. (B) Run-level distributions of ARI decrease for the six leading contributors, illustrating variability across the evaluated analytical pipelines. FSH, anti-thyroglobulin antibodies (anti-TG), anti-thyroid peroxidase antibodies (anti-TPO), total testosterone, LH, and androstenedione produced the largest reductions in phenotype agreement after removal. Thes results reflect contributions to computational robustness and should not be interpreted as measures of causal, diagnostic, or biological importance*.

The strongest effects were observed for follicle-stimulating hormone (FSH), anti-thyroglobulin antibodies (anti-TG), anti-thyroid peroxidase antibodies (anti-TPO), total testosterone, luteinizing hormone (LH), and androstenedione. Removal of these variables produced the largest reductions in phenotype agreement, indicating that they contributed most consistently to the stability of the inferred phenotype structure across alternative analytical workflows.

The feature-level results reflected contributions from more than one biological domain. Gonadotropins and androgen-related measurements were accompanied by thyroid autoimmunity markers among the highest-ranked variables, suggesting that computational phenotype stability was supported by a multidimensional combination of endocrine and immunological information rather than by a single dominant biological axis.

Importantly, the feature perturbation ranking should not be interpreted as a hierarchy of biomarker relevance, disease causality, or diagnostic utility. A high robustness contribution indicates that the presence of a variable helps preserve reproducible patient partitioning under the evaluated analytical perturbations. Conversely, features producing limited changes after removal may still be biologically informative but provide less support for the stability of the unsupervised phenotype solution.

Overall, these analyses identified a restricted set of biomarkers contributing most to computational robustness. The results demonstrate that phenotype reproducibility depends not only on analytical design choices but also on the composition of the feature space used for unsupervised discovery.

### 3.7 Biological validation supports robustness-derived phenotypes

To evaluate whether computational robustness was associated with biological coherence, we compared phenotype solutions exhibiting high robustness with those displaying lower robustness using independent clinical and biochemical characteristics (Figure 7). This analysis was designed as a validation of the robustness framework rather than as a strategy for identifying biologically meaningful phenotypes.

**Figure 7.**
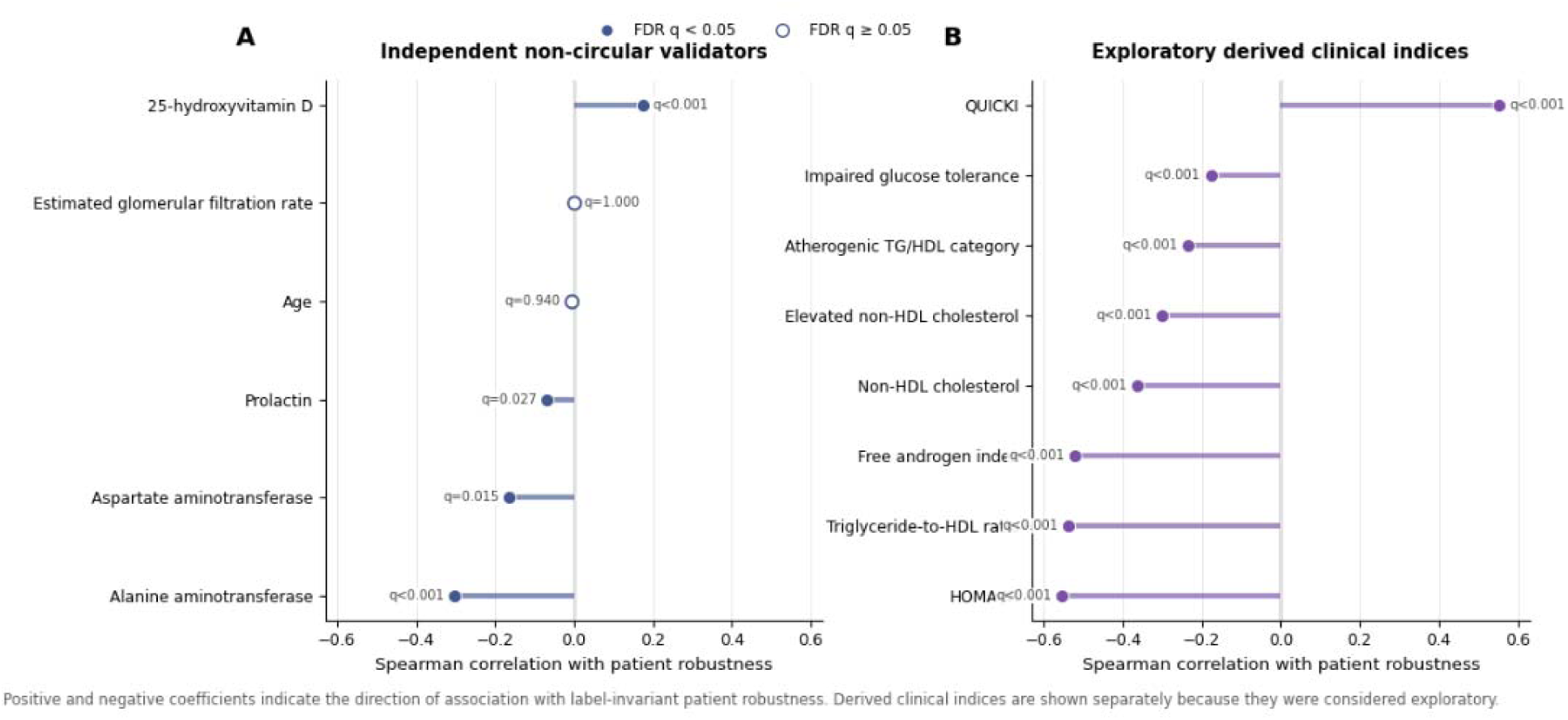
Biological validation of robustness-derived phenotype solutions.

Phenotype solutions classified as highly robust demonstrated consistently greater biological coherence across the evaluated endocrine and metabolic variables. In contrast, phenotype solutions characterized by lower computational robustness showed less distinct biological separation and greater overlap between phenotype groups. These observations indicate that computational reproducibility is accompanied by improved biological consistency.

Importantly, biological coherence was not used during phenotype construction or robustness estimation. Consequently, the observed association represents an independent validation of the analytical framework rather than circular confirmation of the clustering procedure. The robustness metrics therefore provide information that is complementary to conventional biological characterization.

Overall, these findings support the central premise of the proposed framework: phenotype solutions that remain reproducible under realistic analytical perturbations also exhibit greater biological consistency. Rather than defining biological relevance directly, robustness analysis identifies phenotype assignments that are more likely to represent stable and interpretable biological structures.

## 4. Discussion

### 4.1 Principal findings

The present study introduces an analytical perturbation framework for evaluating the robustness of unsupervised phenotype discovery. Although unsupervised clustering has become a central methodology for identifying clinically and biologically meaningful patient subgroups across genomics, transcriptomics, imaging, and electronic health records, the robustness of the resulting phenotype assignments is rarely assessed systematically despite increasing concerns regarding the reproducibility of computational analyses [1–4].

The principal contribution of this work is a shift in perspective from evaluating the performance of individual clustering algorithms to evaluating the stability of the entire analytical workflow. Rather than assuming that a single computational pipeline provides an objective representation of biological heterogeneity, we treated analytically reasonable methodological decisions—including missing-data handling, preprocessing, dimensionality reduction, clustering strategy, and stochastic initialization—as experimental perturbations whose cumulative influence on phenotype discovery can be quantified. This workflow-oriented perspective extends current concepts of clustering stability, which have traditionally focused on algorithmic comparisons, bootstrap resampling, or perturbations of the input data rather than the complete computational pipeline [1,5].

Three principal findings emerge from the present study. First, biologically derived phenotype assignments exhibited substantial instability despite being generated from an identical quality-controlled patient cohort, indicating that analytically plausible methodological choices frequently altered patient classification. Second, this instability originated predominantly from deterministic analytical decisions, particularly preprocessing and clustering strategy, whereas stochastic optimization contributed comparatively little. Finally, phenotype solutions that remained reproducible across analytical perturbations demonstrated greater biological coherence during independent validation, suggesting that computational robustness captures an important property of reliable phenotype discovery rather than representing merely a technical characteristic of the analytical workflow.

These findings have broader implications for computational phenotyping. Modern phenotype discovery increasingly relies on complex multi-step analytical pipelines in which individual methodological choices interact in a non-linear manner. Consequently, evaluating isolated analytical components provides only a partial assessment of reproducibility. By treating the complete computational workflow as the experimental unit, the proposed framework offers a complementary strategy for estimating confidence in unsupervised phenotype assignments and for distinguishing analytically stable structures from those that depend strongly on specific methodological configurations. Such an approach is consistent with the growing emphasis on reproducibility, transparent analytical pipelines, and rigorous computational validation throughout biomedical data science [2–4,6].

Importantly, the proposed framework does not seek to identify a single “true” biological phenotype. Instead, it estimates the degree to which phenotype assignments remain reproducible under analytically reasonable alternative workflows. This distinction is particularly relevant because unsupervised phenotyping is inherently exploratory and lacks an external ground truth against which discovered phenotypes can be validated directly. Rather than replacing conventional biological interpretation, robustness analysis provides an additional quantitative layer of evidence that complements biological characterization and supports more cautious interpretation of computationally derived disease subgroups.

Collectively, our findings suggest that robustness assessment should become an integral component of future computational phenotyping studies. Instead of asking only whether a clustering algorithm identifies biologically meaningful phenotypes, studies should first establish whether those phenotypes remain reproducible under realistic analytical perturbations. We therefore propose analytical robustness as a complementary criterion for interpreting, comparing, and translating computational phenotypes into biologically meaningful disease stratifications.

### 4.2 Why instability arises

The present findings suggest that analytical instability is not an unexpected consequence of imperfect clustering algorithms but rather an intrinsic property of multistep computational phenotyping workflows. Unsupervised learning aims to identify latent structure in high-dimensional data without access to external labels or ground truth. Consequently, the discovered phenotype structure depends on a sequence of analytical decisions that progressively transform the geometry of the data before clustering is performed [1,7]. Each methodological step modifies the representation of relationships between patients and therefore influences the landscape on which clustering algorithms operate.

Among all evaluated analytical components, preprocessing exerted the largest influence on phenotype reproducibility. This observation is consistent with theoretical principles of multivariate analysis, in which feature scaling and normalization directly determine inter-sample distances, covariance structure, and the relative contribution of individual variables to downstream analyses [7–8]. Since most clustering algorithms rely either explicitly or implicitly on distance relationships, modifications introduced during preprocessing propagate throughout the entire analytical workflow and may substantially alter the resulting phenotype partitions. Similar observations have recently been reported in single-cell transcriptomics, where preprocessing decisions frequently produce greater variability in clustering outcomes than the subsequent choice of clustering algorithm itself.

The substantial contribution of the clustering algorithm observed in our study further illustrates that different optimization strategies emphasize distinct geometric properties of the same dataset. Partition-based, hierarchical, and probabilistic clustering algorithms optimize different objective functions and make different assumptions regarding cluster shape, density, and covariance structure [7, 9]. Consequently, when the underlying biological structure is continuous or partially overlapping rather than discretely separated, alternative clustering methods may identify equally plausible yet non-identical phenotype solutions. This phenomenon should not necessarily be interpreted as algorithmic failure but rather as evidence that multiple statistically valid representations of biological heterogeneity may coexist.

In contrast, random initialization explained only a minor proportion of phenotype variability. This finding indicates that the dominant source of instability was systematic methodological variation rather than stochastic optimization. Although random initialization has traditionally been recognized as a potential source of variability in algorithms such as *k*-means, modern implementations generally achieve relatively stable solutions once preprocessing, feature representation, and optimization criteria have been defined [1,9]. Our results therefore suggest that repeated execution of a single analytical pipeline with different random seeds is insufficient for evaluating phenotype robustness because it captures only a small fraction of the uncertainty introduced during computational phenotyping.

Collectively, these observations support a broader conceptual interpretation of computational phenotyping. Rather than viewing preprocessing, dimensionality reduction, clustering, and missing-data handling as independent technical choices, they should be considered interacting components of a unified analytical system. Robust phenotype discovery therefore requires evaluation of the entire analytical workflow, since the cumulative effect of individually reasonable methodological decisions may exceed the influence of any single algorithm. This systems-level perspective extends current approaches to cluster validation and aligns with the increasing emphasis on reproducibility and workflow-aware analyses throughout computational biology and bioinformatics [1–2,6].

### 4.3 Relation to previous literature

The rapid expansion of computational phenotyping has led to widespread adoption of unsupervised learning for identifying disease subtypes across diverse biomedical domains, including transcriptomics, genomics, imaging, and electronic health records [3,7,10]. In most studies, clustering is treated as the final analytical objective, with emphasis placed on selecting an optimal clustering algorithm, tuning model parameters, or comparing internal validity indices. Consequently, biological interpretation is typically based on a single computational solution, while the influence of alternative yet equally plausible analytical workflows remains largely unexplored [3,8].

Several methodological studies have recognized clustering stability as an important indicator of unsupervised learning quality and have proposed validation strategies based on bootstrap resampling, consensus clustering, or perturbation of the input data [1,5]. These approaches provide valuable information regarding algorithmic consistency but generally evaluate only isolated components of the analytical process. In contrast, contemporary computational phenotyping involves a sequence of interdependent methodological decisions—including preprocessing, feature transformation, dimensionality reduction, and clustering—that collectively determine the final phenotype structure. The cumulative influence of these decisions has received comparatively little attention despite increasing awareness that reproducibility should be evaluated at the level of complete computational workflows rather than individual algorithms alone [2,11].

Recent developments in bioinformatics further support this broader perspective. Large-scale benchmarking studies have demonstrated that analytical choices made during data processing frequently exert effects comparable to, or greater than, those of the downstream statistical models themselves [8]. Likewise, reproducibility has emerged as a central theme across computational biology, with recent reviews emphasizing that robust scientific conclusions depend not only on algorithmic performance but also on transparent, reusable, and systematically validated analytical pipelines [2,11]. Our findings are fully consistent with these observations and extend them to the specific context of unsupervised phenotype discovery by demonstrating that methodological perturbations can substantially alter patient-level phenotype assignments even when the underlying dataset remains unchanged.

Our results also complement emerging studies that explicitly evaluate robustness during computational phenotyping. For example, robust clustering frameworks have recently been proposed for multiplexed tissue imaging and other high-dimensional biological datasets, highlighting the importance of reproducible phenotype identification under alternative analytical settings [12]. However, these approaches typically optimize robustness within a specific analytical pipeline or application domain. In contrast, our framework evaluates robustness across multiple complete analytical workflows, enabling uncertainty to be quantified simultaneously at the levels of analytical pipelines, individual patients, and individual features. This workflow-centric perspective represents a conceptual extension of existing stability analyses and provides a more comprehensive characterization of uncertainty in computational phenotype discovery.

Taken together, the present study contributes to an emerging shift within bioinformatics from evaluating individual algorithms toward evaluating the reproducibility of complete computational workflows. Rather than asking which clustering algorithm performs best for a particular dataset, our results suggest that future studies should consider how sensitive biological conclusions are to the sequence of analytical decisions that precede clustering. We therefore view robustness assessment not as an additional validation step performed after phenotype discovery, but as an integral component of the phenotype discovery process itself.

### 4.4 Patient robustness as a new concept

One of the most important conceptual contributions of the proposed framework is the introduction of robustness assessment at the level of individual patients. Existing studies of unsupervised phenotyping generally report cluster-level characteristics, including measures of internal validity, cluster separation, or biological enrichment, while implicitly assuming that every patient assigned to a given cluster belongs to that phenotype with comparable confidence [1,3,5]. In practice, however, patients differ substantially in the stability of their phenotype assignments because some occupy well-defined regions of the multidimensional feature space whereas others lie close to boundaries between alternative phenotype structures.

The Patient Robustness Score proposed in this study addresses this limitation by explicitly quantifying the reproducibility of phenotype assignment across analytically plausible workflows. Rather than asking which phenotype a patient belongs to, the framework estimates how consistently that patient is assigned to the same phenotype despite variation in analytical methodology. This distinction is particularly relevant for exploratory computational phenotyping, where phenotype labels are inferred rather than observed and therefore inherently uncertain [3,7].

From a methodological perspective, the PRS should not be interpreted as a probability of disease membership or diagnostic confidence. Instead, it represents a measure of computational reproducibility, reflecting the stability of phenotype assignment under alternative yet methodologically valid analytical decisions. Consequently, patients with high robustness scores are not necessarily biologically “more typical”; rather, they exhibit phenotype assignments that remain consistent despite changes in preprocessing, dimensionality reduction, clustering strategy, or missing-data handling. Conversely, patients with lower robustness scores may occupy transitional regions of the biological feature space where multiple phenotype representations are statistically plausible.

This interpretation aligns with the increasing recognition that many complex diseases are characterized by continuous biological variation rather than sharply separated subgroups [3, 13–14]. Disease heterogeneity is increasingly viewed as a spectrum in which patients may share overlapping molecular, endocrine, or clinical characteristics rather than belonging to mutually exclusive categories. Within such settings, analytical uncertainty is expected to concentrate among individuals located near phenotype boundaries. The PRS therefore provides an intuitive framework for identifying patients whose computational phenotype assignments should be interpreted with greater caution, without implying that such assignments are incorrect.

Beyond the present application, patient-level robustness may have broader implications for computational medicine. Robustness estimates could complement conventional clustering outputs by providing an additional confidence metric for downstream biological interpretation, hypothesis generation, and external validation. More generally, reporting individual-level robustness may improve transparency and reproducibility in computational phenotyping by explicitly acknowledging that confidence in phenotype assignment is heterogeneous across patients rather than uniform within clusters. We therefore propose patient-level robustness as a complementary dimension of computational phenotyping that extends conventional cluster-based validation toward individual-level assessment of analytical reliability.

### 4.5 Computational robustness versus biological importance

An important distinction emerging from the present study concerns the interpretation of feature-level robustness. The perturbation analyses identified a relatively small group of variables— including FSH, anti-TG, anti-TPO, total testosterone, and LH—that contributed most strongly to the stability of phenotype discovery across alternative analytical workflows. However, these findings should not be interpreted as evidence that these biomarkers are biologically more important, causally related to disease pathogenesis, or diagnostically superior to other variables. Instead, they identify biomarkers that contribute most consistently to the computational robustness of the inferred phenotype structure.

This distinction is particularly important in biomedical machine learning, where feature rankings are frequently interpreted as measures of biological relevance. Numerous studies have demonstrated that variables identified as highly influential by machine learning models do not necessarily correspond to causal mechanisms or clinically dominant biomarkers, but instead reflect their contribution to prediction, discrimination, or model stability [15–18]. Consequently, computational importance and biological importance represent related but fundamentally different concepts that should not be considered interchangeable.

Within the proposed framework, feature robustness quantifies the extent to which removal of an individual variable disrupts reproducible phenotype assignment across analytically plausible workflows. Variables receiving high robustness scores therefore act as stabilizing components of the computational representation rather than as direct indicators of disease biology. Conversely, biomarkers producing relatively small perturbation effects may still play important biological or clinical roles but contribute less to maintaining a stable computational partition of the patient population. This interpretation is analogous to observations in explainable artificial intelligence, where variables with the greatest influence on model behaviour are not necessarily those with the strongest biological or causal significance [16–17].

The presence of endocrine and thyroid-related biomarkers among the strongest contributors to computational robustness is nevertheless biologically plausible. PCOS is increasingly recognized as a multidimensional disorder involving interactions between reproductive endocrinology, metabolism, and immune regulation rather than a single dominant pathological pathway [19–20]. It is therefore reasonable that biomarkers representing complementary physiological domains contribute disproportionately to maintaining a reproducible phenotype structure. Importantly, our framework does not identify these variables because they exhibit the largest effect sizes or strongest statistical associations, but because they consistently preserve the stability of phenotype assignments across alternative analytical workflows.

More broadly, distinguishing computational robustness from biological importance may help avoid a common source of overinterpretation in computational phenotyping studies. Feature rankings derived from clustering, prediction, or explainability analyses are often discussed as if they directly reflected disease mechanisms. Our results suggest a more cautious interpretation: computational robustness provides information about the structural stability of phenotype discovery, whereas biological interpretation requires independent experimental, clinical, or mechanistic evidence. We therefore view feature robustness as a complementary analytical property that enhances confidence in computational phenotypes without replacing conventional biological validation.

### 4.6 Implications

The implications of the proposed framework extend beyond the specific application presented in this study. Although PCOS served as an appropriate use case because of its well-recognized biological heterogeneity, the analytical perturbation framework was designed as a general methodology for evaluating the robustness of unsupervised phenotype discovery. Consequently, its underlying principles are applicable to any computational phenotyping task in which patient subgroups are inferred from high-dimensional biomedical data rather than directly observed.

This perspective is particularly relevant as biomedical research increasingly relies on unsupervised analyses of large-scale multimodal datasets, including transcriptomics, proteomics, metabolomics, medical imaging, digital phenotyping, and electronic health records [3, 10, 21]. Across these domains, computational phenotypes are frequently interpreted as biologically meaningful entities despite substantial variability introduced by analytical workflow design. Our findings suggest that reporting a single clustering solution without quantifying its robustness may provide an incomplete representation of the underlying biological structure. Instead, robustness assessment should accompany phenotype discovery in the same way that uncertainty estimation accompanies statistical inference.

The proposed framework also aligns with the broader goals of precision medicine. Patient stratification increasingly forms the basis for disease subclassification, biomarker discovery, treatment selection, and risk prediction [14, 22]. However, if phenotype assignments depend strongly on analytical decisions, downstream clinical interpretation may also become workflow-dependent. Quantifying robustness before biological interpretation therefore provides an additional safeguard against overinterpreting computationally unstable patient subgroups and may improve the reproducibility of translational research.

Beyond evaluating analytical workflows, the proposed framework introduces a complementary paradigm for reporting computational phenotyping studies. Rather than presenting phenotype assignments as deterministic outputs, future studies may report both phenotype membership and an accompanying robustness estimate at the level of individual patients and discovered phenotypes. Such an approach acknowledges the inherent uncertainty of unsupervised learning while preserving the interpretability and practical utility of computational phenotypes.

Although developed in the context of clinical phenotyping, the framework is readily transferable to other bioinformatics applications involving unsupervised structure discovery, including molecular subtype identification, single-cell analyses, spatial transcriptomics, microbiome profiling, and multi-omics integration [10, 21, 23]. As increasingly complex analytical pipelines become standard practice in biomedical research, evaluating robustness at the level of the complete computational workflow may become an essential component of reproducible data-driven discovery.

Collectively, these considerations suggest that robustness assessment should evolve from a supplementary validation procedure into a routine element of computational phenotyping. We envision analytical robustness not as an additional performance metric, but as a fundamental descriptor of confidence that complements biological interpretation and supports the reproducible translation of computational phenotypes into biomedical research and precision medicine.

### 4.7 Strengths and limitations

The present study possesses several methodological strengths that distinguish it from conventional computational phenotyping analyses. First, robustness was evaluated across complete analytical workflows rather than isolated clustering algorithms, thereby reflecting the cumulative influence of realistic methodological decisions encountered during biomedical data analysis. Second, robustness was quantified simultaneously at multiple complementary levels— including analytical pipelines, individual patients, and individual features—providing a comprehensive assessment of computational uncertainty that extends beyond conventional cluster validation metrics. Third, biological characterization was performed independently of the robustness estimation procedure, reducing the risk of circular interpretation and allowing biological coherence to serve as an external validation of the proposed framework. Together, these features provide a reproducible strategy for evaluating computational phenotype discovery that is readily transferable to other biomedical applications.

Several limitations should nevertheless be considered when interpreting the present findings. First, the framework was evaluated using a single-center cohort of women with PCOS. Although this dataset represents a clinically well-characterized and biologically heterogeneous population that is well suited for methodological evaluation, additional studies involving independent cohorts, different disease entities, and alternative data modalities will be required to establish the generalizability of the proposed framework. Future validation across external datasets will be particularly important for determining how robustness behaves in populations differing in demographic composition, clinical practice, and data acquisition protocols.

Second, the analytical perturbation space was intentionally restricted to methodological decisions commonly encountered during unsupervised phenotype discovery. While these choices encompass major sources of analytical variability—including missing-data handling, preprocessing, dimensionality reduction, clustering methodology, and stochastic initialization— they do not exhaust all possible sources of uncertainty. Future extensions of the framework could incorporate additional perturbations, such as alternative feature selection strategies, nonlinear representation learning, deep clustering approaches, graph-based methods, or multimodal data integration, thereby providing an even broader characterization of computational robustness.

Finally, although robustness was associated with greater biological coherence, computational reproducibility should not be interpreted as evidence of biological truth. Robust phenotype assignments represent solutions that remain stable across analytically plausible workflows, but their biological relevance must continue to be established through independent clinical studies, mechanistic investigations, functional experiments, and prospective validation. Accordingly, robustness should be viewed as a complementary criterion that increases confidence in computational phenotypes without replacing biological or clinical validation [2–3,21].

Despite these limitations, we believe that the proposed framework provides an important step toward more transparent and reproducible computational phenotyping. As biomedical datasets continue to increase in size, complexity, and dimensionality, evaluating the robustness of analytical workflows will become increasingly important for ensuring that discovered phenotypes reflect stable computational structures rather than artifacts of methodological choice. We therefore anticipate that robustness assessment will evolve from an optional validation procedure into a routine component of unsupervised phenotype discovery and, more broadly, reproducible biomedical data science.

## 5. Conclusions

We present an analytical perturbation framework for evaluating the robustness of unsupervised phenotype discovery across complete computational workflows. By systematically perturbing preprocessing, missing-data handling, dimensionality reduction, clustering methodology, and stochastic initialization, the framework quantifies the reproducibility of phenotype assignments at the levels of analytical pipelines, individual patients, and individual features. This workflow-oriented perspective extends conventional approaches to cluster validation by explicitly incorporating analytical uncertainty into computational phenotyping.

Our findings demonstrate that computational phenotype discovery is substantially influenced by methodological decisions that are often regarded as routine components of the analytical workflow. In particular, deterministic analytical choices contributed far more to phenotype variability than stochastic optimization, while independent biological validation showed that phenotype solutions exhibiting greater computational robustness also displayed greater biological coherence. These observations suggest that robustness assessment provides complementary information that cannot be obtained from conventional cluster validity metrics alone.

Importantly, the proposed framework does not attempt to identify a single “true” biological phenotype. Instead, it provides a quantitative estimate of the confidence with which computational phenotypes remain reproducible under analytically plausible alternative workflows. This distinction is particularly relevant for exploratory analyses of complex biomedical datasets, where multiple statistically valid representations of biological heterogeneity may coexist.

We therefore propose that robustness assessment should become a routine component of computational phenotyping studies. Rather than interpreting a single clustering solution as definitive, future analyses should evaluate whether discovered phenotypes remain stable across realistic analytical perturbations. We anticipate that this framework will facilitate more transparent, reproducible, and biologically interpretable phenotype discovery across a broad range of biomedical applications, including multi-omics integration, single-cell analysis, digital phenotyping, and precision medicine.

## Data Availability

The clinical dataset analyzed during the current study is not publicly available because it contains sensitive patient information and institutional ethical restrictions prohibit unrestricted public release. De-identified data may be made available from the corresponding author upon reasonable request and subject to approval by the appropriate institutional and ethical authorities.

## Code Availability

The complete analytical workflow developed for this study, including Python scripts for data harmonization, preprocessing, missing-data handling, dimensionality reduction, clustering, stability assessment, cross-space agreement analysis, biological validation, sensitivity analyses, and figure generation, is openly available in the following GitHub repository: https://github.com/npiorkowska-science/

The repository also contains documentation describing the computational workflow, software dependencies, and instructions required to reproduce all analyses presented in this manuscript.

## Ethics Approval

This study was conducted in accordance with the Declaration of Helsinki and was approved by the Bioethics Committee of Wroclaw Medical University (approval no. 254/2021).

## Consent to Participate

Written informed consent was obtained from all participants prior to inclusion in the study.

## Consent for Publication

All authors have reviewed and approved the final version of the manuscript and consent to its publication.

## Author Contributions

Natalia Piórkowska: Conceptualization, methodology, software, formal analysis, investigation, data curation, visualization, writing – original draft, writing – review and editing.

Alan Ostromęcki: Methodology, software validation, formal validation, writing – review and editing.

Grzegorz Franik: Data curation.

Anna Bizoń: Writing – review and editing, supervision. All authors read and approved the final manuscript.

## Acknowledgements

The authors thank all study participants and the clinical staff involved in patient recruitment, data collection, and laboratory diagnostics.

## Tables

**Table 1**. Characteristics of the study cohort and analytical perturbation framework.

## Figures

**Figure 1**. Analytical Perturbation Framework for robustness assessment of unsupervised biological phenotypes.

**Figure 2**. Global robustness of phenotype assignments across analytical perturbations.

**Figure 3**. Pairwise agreement between phenotype assignments generated by alternative analytical workflows.

**Figure 4**. Relative contribution of analytical decisions to phenotype instability.

**Figure 5**. Distribution of patient-level phenotype robustness.

**Figure 6**. Feature-level contributions to the robustness of phenotype discovery.

**Figure 7**. Biological validation of robustness-derived phenotype solutions.

## References

[1] Liu T, Yu H, Blair RH. Stability estimation for unsupervised clustering: a review. WIREs Computational Statistics. 2022.

[2] Baykal PI et al. Genomic reproducibility in the bioinformatics era. Genome Biology. 2024.

[3] He T et al. Trends and opportunities in computable clinical phenotyping: a scoping review. Journal of Biomedical Informatics. 2023.

[4] Ding S et al. Identify and mitigate bias in electronic phenotyping: a computational perspective. Journal of Biomedical Informatics. 2024.

[5] von Luxburg U. Clustering stability: an overview. Foundations and Trends in Machine Learning. 2010;2(3):235–274.

[6] Sandve GK, Nekrutenko A, Taylor J, Hovig E. Ten Simple Rules for Reproducible Computational Research. PLoS Computational Biology. 2013;9(10):e1003285.

[7] Neijzen D, et al. Unsupervised learning for medical data: A review of probabilistic factorization methods. Statistics in Medicine. 2023.

[8] Ospina O, Soupir A, Fridley BL. A Primer on Preprocessing, Visualization, Clustering, and Phenotyping of Barcode-Based Spatial Transcriptomics Data. Methods in Molecular Biology. 2023.

[9] Jaeger A, Banks D. Cluster analysis: A modern statistical review. WIREs Computational Statistics. 2023.

[10] Brooks TG, Lahens NF, Mrčela A, et al. Challenges and best practices in omics benchmarking. Nature Reviews Genetics. 2024.

[11] Alnasir J, et al. Developing and reusing bioinformatics data analysis pipelines using scientific workflow systems. Computational and Structural Biotechnology Journal. 2023.

[12] Liu CC, Greenwald N, Kong A, et al. Robust phenotyping of highly multiplexed tissue imaging data using pixel-level clustering. Nature Communications. 2023.

[13] Robinson PN. Deep phenotyping for precision medicine. Human Mutation. 2012. (klasyczna praca definiująca ideę głębokiego fenotypowania)

[14] Köhler S, Gargano M, Matentzoglu N, et al.; Robinson PN. The Human Phenotype Ontology in 2021. Nucleic Acids Research. 2021;49(D1):D1207–D1217.

[15] Molnar C. Interpretable Machine Learning. 2nd edition, 2022. (rozdziały dotyczące interpretacji ważności cech).

[16] Rudin C. Stop explaining black box machine learning models for high stakes decisions and use interpretable models instead. Nature Machine Intelligence. 2019.

[17] Lundberg SM, Lee SI. A Unified Approach to Interpreting Model Predictions. NeurIPS. 2017. (lub nowszy przegląd dotyczący SHAP i interpretowalności w biomedycynie, jeśli chcemy ograniczyć starsze cytowania).

[18] Biecek P, Burzykowski T. Explanatory Model Analysis. CRC Press. 2021. *(lub nowszy artykuł przegl*ą*dowy dotycz*ą*cy interpretowalno*ś*ci modeli ML)*.

[19] International Evidence-based Guideline for the Assessment and Management of Polycystic Ovary Syndrome. 2023.

[20] Azziz R, Carmina E, Dewailly D, et al. (incl. Legro RS). Polycystic ovary syndrome. Nature Reviews Disease Primers. 2016;2:16057.

[21] Hasin Y, Seldin M, Lusis A. Multi-omics approaches to disease. Genome Biology. 2017;18:83.

[22] Collins FS, Varmus H. A New Initiative on Precision Medicine. New England Journal of Medicine, 2015;372(9):793–795.

[23] Lähnemann D, Köster J, Szczurek E, et al. Eleven grand challenges in single-cell data science. Genome Biology. 2020 21:31.

